# Metabolic and muscle-derived serum biomarkers define CHCHD10-linked late-onset spinal muscular atrophy

**DOI:** 10.1101/2021.04.07.21254960

**Authors:** Julius Järvilehto, Sandra Harjuhaahto, Edouard Palu, Mari Auranen, Jouni Kvist, Henrik Zetterberg, Johanna Koskivuori, Marko Lehtonen, Anna Maija Saukkonen, Manu Jokela, Emil Ylikallio, Henna Tyynismaa

**Affiliations:** Stem Cells and Metabolism Research Program, Faculty of Medicine, University of Helsinki, Helsinki, Finland; Clinical Neurosciences, Neurology, Helsinki University Hospital, Helsinki, Finland; Clinical Neurochemistry Laboratory, Sahlgrenska University Hospital, Mölndal, Sweden; Department of Psychiatry and Neurochemistry, Institute of Neuroscience and Physiology, the Sahlgrenska Academy at the University of Gothenburg, Mölndal, Sweden; Department of Neurodegenerative Disease, UCL Institute of Neurology, London, United Kingdom; UK Dementia Research Institute at UCL, London, United Kingdom; School of Pharmacy, University of Eastern Finland, Kuopio, Finland; Department of Neurology, Central Hospital of Northern Karelia, Joensuu, Finland; Division of Clinical Neurosciences, Turku University Hospital and University of Turku, Turku, Finland; Department of Neurology, Neuromuscular Research Center, Tampere University Hospital and Tampere University, Tampere, Finland; Neuroscience Center, Helsinki Institute of Life Science, University of Helsinki, Helsinki, Finland; Department of Medical and Clinical Genetics, University of Helsinki, Helsinki, Finland

## Abstract

**Objective:** To characterize serum biomarkers in mitochondrial CHCHD10-linked spinal muscular atrophy Jokela type (SMAJ) for disease monitoring and for understanding of pathogenic mechanisms.

**Methods:** We collected serum samples from a cohort of 49 SMAJ patients, all carriers of the heterozygous c.197G>T p.G66V variant in *CHCHD10*. As controls, we used age- and sex-matched serum samples obtained from Helsinki Biobank. Neurofilament light (NfL) and glial fibrillary acidic protein (GFAP) were measured with Single molecule array (Simoa), and fibroblast growth factor 21 (FGF-21) and growth differentiation factor 15 (GDF-15) with enzyme-linked immunosorbent assay. For nontargeted serum metabolite profiling, samples were analyzed with liquid chromatography–high resolution mass spectrometry. Disease severity was evaluated retrospectively by calculating a symptom-based score.

**Results:** Axon degeneration marker NfL was unexpectedly not altered in the serum of SMAJ patients, whereas astrocytic activation marker GFAP was slightly decreased. Creatine kinase was elevated in most patients, in particular males. We identified six metabolites that were significantly altered in SMAJ patients’ serum compared to controls: increased creatine and pyruvate, and decreased creatinine, taurine, N-acetyl-carnosine and succinate. Creatine correlated with disease severity. Altered pyruvate and succinate indicated a metabolic response to mitochondrial dysfunction, however, lactate or mitochondrial myopathy markers FGF-21 or GDF-15 were not changed.

**Conclusions:** Biomarkers of muscle mass and damage are altered in SMAJ serum, indicating a role for skeletal muscle in disease pathogenesis in addition to neurogenic damage. Despite the minimal mitochondrial pathology in skeletal muscle, signs of a metabolic shift can be detected in the serum of SMAJ patients.

## INTRODUCTION

Spinal muscular atrophy Jokela type (SMAJ, MIM#615048) is a progressive adult-onset lower motor neuron disease (1). Muscle cramps are a common initial manifestation in SMAJ, typically appearing after age 30-40 (2). Increasing lower limb predominant muscle weakness, hyporeflexia and difficulties in walking follow as the disease progresses. Mild distal sensory defects occur in about half of the patients later in the disease course (2). Life expectancy is within normal range (2). SMAJ patients have normal nerve conduction velocities but EMG shows widespread neurogenic alterations (1,3). Muscle biopsies also show characteristic neurogenic findings, including fiber type grouping and group atrophy, whereas only some patients have mild signs of mitochondrial myopathy such as a few COX-negative muscle fibers (3). Elevated creatine kinase (CK) has been reported in SMAJ (1,2). Some patients have received a clinical diagnosis of amyotrophic lateral sclerosis (ALS) or Charcot-Marie-Tooth (CMT) neuropathy (4,5,6), but a gene test of a single heterozygous variant, c.197G>T p.G66V, in *coiled-coil-helix-coiled-coil-helix domain containing 10* (*CHCHD10*) sets the diagnosis of SMAJ (2). Other more severe *CHCHD10* variants can cause ALS and frontotemporal dementia (FTD) (5) or mitochondrial myopathy (7).

CHCHD10 is a mitochondrial of unknown exact function, and although it is located in the intermembrane space, both its absence and dominant pathogenic mutations affect mitochondrial respiration (8,9,10,11,12). Recent genetically modified Chchd10 mice have displayed neuromuscular junction (NMJ) and motor neuron degeneration (9,14). Mice with ALS variant p.S59L developed mitochondrial dysfunction in muscle before NMJ degeneration (9). Toxic gain-of-function effect of mutant Chchd10 aggregates were suggested to induce mitochondrial integrated stress response (ISR) (8), which was also reported in fibroblasts of ALS patients with the same mutation (14). Besides fibroblasts, studies using samples from patients with *CHCHD10* mutations have so far been rare.

Currently, no serum-based tests for diagnostic use or for monitoring disease progression are available for SMAJ. Here we present an investigation of serum biomarkers in SMAJ, an adult-onset non-5q spinal muscular atrophy of mitochondrial origin. We evaluate neurofilament light (NfL) and glial fibrillary acidic protein (GFAP) as neuronal injury and astrocytic activation markers, and fibroblast growth factor 21 (FGF-21) and growth differentiation factor 15 (GDF-15) as mitochondrial stress markers, and perform a nontargeted metabolite screen for unbiased biomarker discovery.

## PATIENTS AND METHODS

This study was approved by the medical ethics committee of Helsinki University Hospital (project code U1024NEURO).

In total 49 individuals who were symptomatic, genetically verified carriers of CHCHD10 p.G66V variant were available and recruited for the study. We used a questionnaire and review of medical charts to collect clinical data. Fasting serum samples were collected and stored at −80°C until the analysis. As controls we used anonymized samples from Helsinki Biobank, which were requested to match the sex and age distributions of the study population, but with exclusion of neurological disease diagnosis (ICD-10 code starting with G). The number of samples used in each analyses varied depending on availability at the time of the measurement (Table 1).

**Table 1.** Demographics of the study groups in different analyses.

| Analysis | Patients |  | Controls |  |
| --- | --- | --- | --- | --- |
|  | n (M/F) | Age median (range) | n (M/F) | Age median (range) |
| S-NFL<br>S-GFAP | 49 (20/29) | 65 (39-86) | 45 (17/28) | 64 (40-86) |
| S-FGF-21<br>S-GDF-15 | 49 (20/29) | 65 (39-86) | 24 (10/14) | 63 (47-85) |
| Metabolite profiling | 43 (19/24) | 65 (39-86) | 48 (18/30) | 65 (34-86) |
| S-CK<br>S-Creatinine | 49 (20/29) | 65 (39-86) | -<br>(reference values) | -<br>(reference values) |
| Symptom score | 43 (16/27) | 65 (39-85) | - | - |

Disease severity was determined by using a symptom-based score, which we modified from the motor and sensory defect part of a more comprehensive CMT neuropathy score (15) (Table 2). Sufficient patient records were available for assessing the score for 43 patients. Certain patients’ symptoms were not strong enough to merit positive scores on this scale (e.g. only cramps), and thus received a symptom score of 0. As SMAJ is a progressive disease, the symptom score correlated with age (r=0.56, p<0.001).

**Table 2.** Symptom score for SMAJ patients, modified from the motor and sensory defect part of a more comprehensive CMT-score (10).

| Parameter | 0 | 1 | 2 | 3 | 4 |
| --- | --- | --- | --- | --- | --- |
| Sensory symptoms | None | Symptoms below or at ankle bones | Symptoms up to the distal half of the calf | Symptoms up to the proximal half of the calf, including knee | Symptoms above the knee (above the top of the patella) |
| Motor symptoms (legs) | None | Trips, catches toes, slaps foot, shoe inserts | Ankle support or stabilization (AFOs), foot surgery | Walking aids (cane, walker) | Wheelchair |
| Motor symptoms (arms) | None | Mild difficulty with buttons | Severe difficulty or unable to do buttons | Unable to cut most foods | Proximal weakness, (affect movements involving the elbow and above) |

### Serum measurements

Serum NfL and GFAP concentrations were measured with Single molecule array (Simoa) technology on an HD-X Analyzer using commercially available NF-light Advantage and GFAP Discovery kits (Quanterix, Billerica, MA). Serum FGF-21 and GDF-15 levels were measured with R&D Systems Quantikine ELISA kits (DF2100 and DGD150, respectively; R&D Systems, Minneapolis, MN). Creatinine and CK were measured in accredited laboratory HUSLAB using a photometric enzymatic method.

### Nontargeted metabolite profiling

Nontargeted metabolite profiling on serum samples was performed at the LC-MS metabolomics center (University of Eastern Finland, Kuopio) using ultra-high performance liquid chromatography coupled to high-resolution mass spectrometry. Before the analysis, samples were randomized and deproteinized with acetonitrile. A small volume of each sample was pooled for quality control (QC) sample and were analyzed at the beginning and after every 12^th^ sample of the instrument worklist. To meet the wide diversity of sample components, all samples were analyzed using two different chromatographic techniques; i.e., reversed phase (RP) and hydrophilic interaction chromatography (HILIC). In addition, data were acquired in both mass spectrometric electrospray ionization polarities; i.e., ESI+ and ESI-(16).

### Data processing and statistical analysis

Statistical analysis was performed using GraphPad Prism 8.42 (GraphPad; La Jolla, CA). Patient and control serum NfL, GFAP, FGF-21 and GDF-15 concentrations, as well as metabolite abundances were compared using Mann-Whitney U-test analysis. Correlations were assessed using Spearman correlation coefficients. P-value < 0.05 was considered significant, except in metabolome profiling where adjusted p-value < 0.05 was considered significant.

### Data availability statement

Anonymized data will be shared by request from any qualified investigator.

## RESULTS

### Serum NfL is not elevated in SMAJ

The axonal injury protein NfL has shown to be a potent serum biomarker in a number of neurological conditions (17), including ALS, CMT and SMA (18,19,20). However, in our measurements, the level of serum NfL was not significantly different in SMAJ patient samples (median 14.9 pg/ml) compared to controls (13.6 pg/ml) (Fig 1A). GFAP was also not elevated in SMAJ, in contrast decreased level was observed in SMAJ patients’ serum (median 118 pg/ml) as compared to controls (177 pg/ml) (p = 0.006, Fig 1B). GFAP, a marker of astrogliosis (21), is centrally expressed and an autopsied individual with SMAJ did not have spinal cord abnormalities (4). The levels of NfL and GFAP increased as a function of age in both patient and control groups, but neither marker correlated with the symptom score (Table 3).

**Table 3.** Correlations between different markers

|  | Age | Sex | Symptom score | Creatine | Creatinine | CK | Taurine | N-acetyl-L-carnosine | Pyruvate | Succinate | FGF-21 | GDF-15 | GFAP | NfL |
| --- | --- | --- | --- | --- | --- | --- | --- | --- | --- | --- | --- | --- | --- | --- |
| Creatine | **r = 0.40**<br>**p = 0.009** | p = 0.056 | **r = 0.43**<br>**p = 0.009** |  | r = -0.16<br>p = 0.31 | **r = 0.34**<br>**p = 0.03** | r = 0.27<br>p = 0.08 | r = -0.25<br>p = 0.11 | r = 0.038<br>p = 0.81 | r = 0.17<br>p = 0.27 | **r = 0.32**<br>**p = 0.04** | r = 0.10<br>p = 0.52 | **r = 0.41**<br>**p = 0.008** | r = 0.27<br>p = 0.09 |
| Creatinine | r = 0.22<br>p = 0.16 | p = 0.12 | r = -0.11<br>p = 0.52 | r = -0.16<br>p = 0.31 |  | r = 0.15<br>p = 0.36 | r = 0.015<br>p = 0.92 | **r = 0.60**<br>**p < 0.001** | r = 0.18<br>p = 0.24 | r = 0.0078<br>p = 0.96 | r = 0.0088<br>p = 0.96 | r = 0.014<br>p = 0.93 | r = 0.047<br>p = 0.77 | r = 0.10<br>p = 0.53 |
| CK | r = 0.079<br>p = 0.62 | **p < 0.0001** | r = 0.16<br>p = 0.35 | **r = 0.34**<br>**p = 0.03** | r = 0.15<br>p = 0.36 |  | r = -0.0014<br>p > 0.99 | r = 0.022<br>p = 0.89 | r = -0.10<br>p = 0.52 | r = 0.28<br>p = 0.07 | r = -0.14<br>p = 0.34 | r = -0.10<br>p = 0.48 | r = -0.18<br>p = 0.21 | r = -0.12<br>p = 0.43 |
| Taurine | r = 0.15<br>p = 0.32 | p = 0.17 | r = -0.33<br>p = 0.05 | r = 0.27<br>p = 0.08 | r = 0.015<br>p = 0.92 | r = -0.0014<br>p > 0.99 |  | r = -0.019<br>p = 0.90 | r = -0.19<br>p = 0.23 | **r = 0.37**<br>**p = 0.01** | r = 0.27<br>p = 0.09 | r = -0.0094<br>p = 0.95 | r = 0.22<br>p = 0.17 | r = 0.30<br>p = 0.06 |
| N-acetyl L-carnosine | r = -0.032<br>p = 0.77 | p = 0.35 | r = -0.10<br>p = 0.54 | r = -0.25<br>p = 0.11 | **r = 0.60**<br>**p < 0.001** | r = 0.022<br>p = 0.89 | r = -0.019<br>p = 0.90 |  | r = 0.084<br>p = 0.59 | r = 0.066<br>p = 0.67 | r = -0.083<br>p = 0.61 | r = 0.21<br>p = 0.18 | r = -0.050<br>p = 0.75 | r = -0.12<br>p = 0.44 |
| Pyruvate | r = 0.24<br>p = 0.13 | p = 0.07 | r = 0.28<br>p = 0.10 | r = 0.038<br>p = 0.81 | r = 0.18<br>p = 0.24 | r = -0.10<br>p = 0.52 | r = -0.19<br>p = 0.23 | r = 0.084<br>p = 0.59 |  | r = 0.082<br>p = 0.60 | r = 0.054<br>p = 0.74 | r = 0.072<br>p = 0.65 | r = 0.14<br>p = 0.37 | r = 0.018<br>p = 0.91 |
| Succinate | r = 0.22<br>p = 0.16 | p = 0.86 | r = -0.044<br>p = 0.80 | r = 0.17<br>p = 0.27 | r = 0.0078<br>p = 0.96 | r = 0.28<br>p = 0.07 | **r = 0.37**<br>**p = 0.01** | r = 0.066<br>p = 0.67 | r = 0.082<br>p = 0.60 |  | r = 0.11<br>p = 0.50 | r = 0.17<br>p = 0.28 | r = 0.046<br>p = 0.77 | r = 0.25<br>p = 0.11 |
| FGF-21 | r = 0.022<br>p = 0.88 | p = 0.96 | r = 0.043<br>p = 0.79 | **r = 0.32**<br>**p = 0.04** | r = 0.0088<br>p = 0.96 | r = -0.14<br>p = 0.34 | r = 0.27<br>p = 0.09 | r = -0.083<br>p = 0.61 | r = 0.054<br>p = 0.74 | r = 0.11<br>p = 0.50 |  | **r = 0.35**<br>**p = 0.02** | **r = 0.32**<br>**p = 0.03** | r = 0.085<br>p = 0.57 |
| GDF-15 | r = 0.18<br>p = 0.21 | p = 0.22 | r = 0.18<br>p = 0.26 | r = 0.10<br>p = 0.52 | r = 0.014<br>p = 0.93 | r = -0.10<br>p = 0.48 | r = -0.0094<br>p = 0.95 | r = 0.21<br>p = 0.18 | r = 0.072<br>p = 0.65 | r = 0.17<br>p = 0.28 | **r = 0.35**<br>**p = 0.02** |  | r = 0.25<br>p = 0.08 | r = 0.084<br>p = 0.56 |
| GFAP | **r = 0.57**<br>**p < 0.001** | p = 0.88 | r = 0.25<br>p = 0.11 | **r = 0.41**<br>**p = 0.008** | r = 0.047<br>p = 0.77 | r = -0.18<br>p = 0.21 | r = 0.22<br>p = 0.17 | r = -0.050<br>p = 0.75 | r = 0.14<br>p = 0.37 | r = 0.046<br>p = 0.77 | **r = 0.32**<br>**p = 0.03** | r = 0.25<br>p = 0.08 |  | **r = 0.59**<br>**p < 0.001** |
| NfL | **r = 0.55**<br>**p < 0.001** | p = 0.37 | r = 0.066<br>p = 0.68 | r = 0.27<br>p = 0.09 | r = 0.10<br>p = 0.53 | r = -0.12<br>p = 0.43 | r = 0.30<br>p = 0.06 | r = -0.12<br>p = 0.44 | r = 0.018<br>p = 0.91 | r = 0.25<br>p = 0.11 | r = 0.085<br>p = 0.57 | r = 0.084<br>p = 0.56 | **r = 0.59**<br>**p < 0.001** |  |

**Figure 1.**
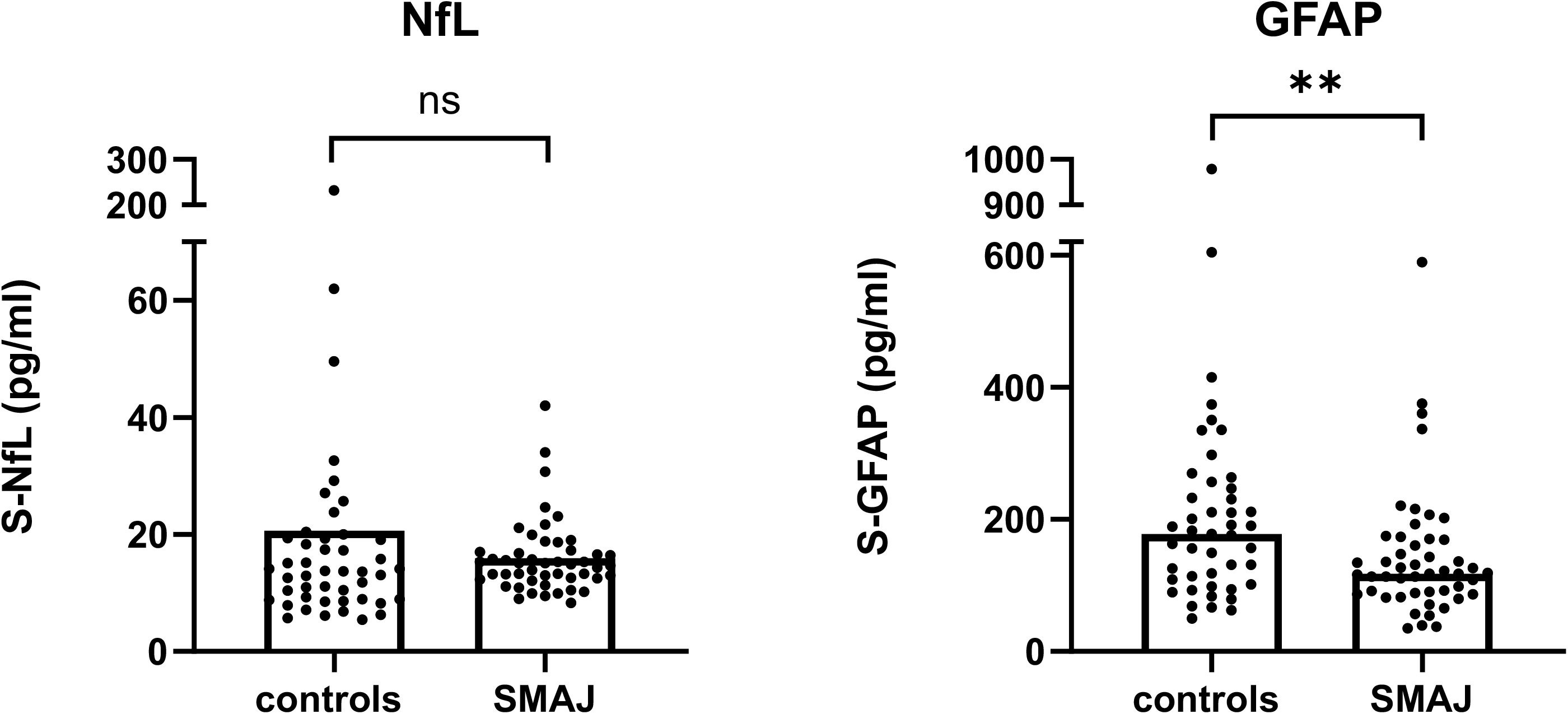
Serum neurofilament light is not elevated in SMAJ. Comparison of serum concentrations of NfL and GFAP between SMAJ patients (n=49) and controls (n=45). Median per group is indicated by bar. ** = p<0.01, Mann-Whitney U-test.

### Serum biomarkers of muscle-origin define SMAJ

Previous studies had indicated that serum CK may be elevated in SMAJ (1,2). We measured CK in serum samples of patients included in this study, and found that indeed CK was markedly higher in SMAJ patients as compared to the reference values (Fig 2A). Of the female SMAJ patients, 60% had CK values above reference (mean 318 U/l, reference range 35-210 U/l in females over 18), whereas nearly all (95%) male SMAJ patients had elevated CK values (mean 711 U/l, reference range 40-280 U/l in males over 50). CK values did not, however, correlate with the symptom score (r = 0.16, p = 0.35) (Fig 2D).

**Figure 2.**
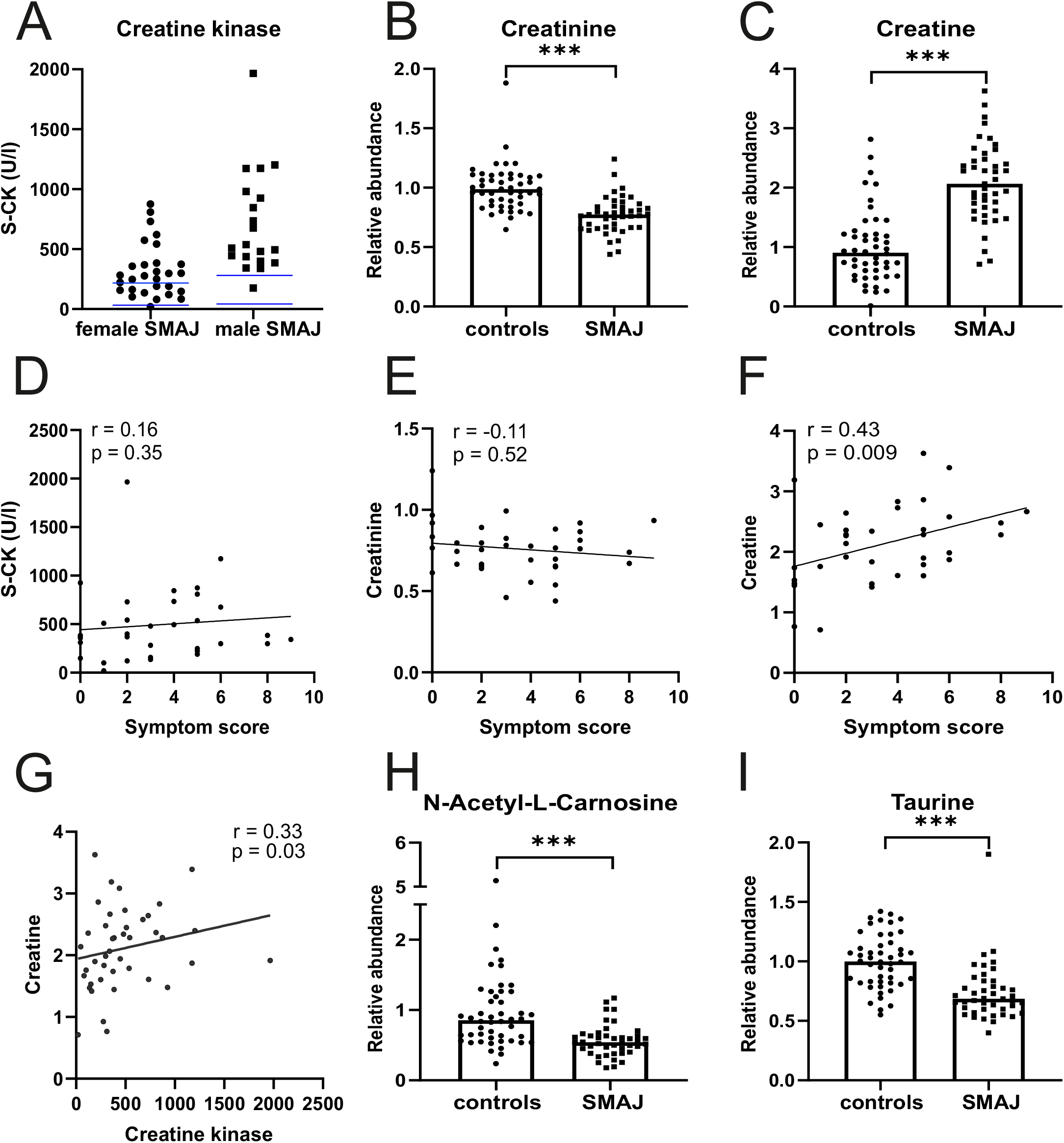
Biomarkers of muscle damage and mass in SMAJ serum. **A)** Creatine kinase (S-CK) units of enzyme activity per liter of serum in female (n=29) and male (n=20) SMAJ patients. Blue lines indicate the upper and lower age- and sex-matched laboratory reference values. Comparison of serum **B)** creatinine and **C)** creatine values between SMAJ patients (n=43) and controls (n=48) measured with mass spectrometer. Correlation of **C)** S-CK, **D)** creatinine and **E)** creatine to symptom score by Spearman correlation test. For all, n = 36. **F)** Correlation of creatine to CK Spearman correlation test n = 43. Comparison of serum **G)** N-acetyl-L-carnosine and **H)** taurine values between SMAJ patients (n=43) and controls (n=48) measured with mass spectrometer. For comparisons median per group is indicated by bar. *** = p< 0.0001, Mann-Whitney U-test.

Next we performed a nontargeted metabolite screen with patient and control serum samples to identify metabolites that could be used as markers of SMAJ pathogenesis. We focused on six metabolites that were significantly altered in SMAJ samples in comparison to controls: creatine (Fold change 2.09, adjusted p-value 6.13E-07), creatinine (FC −1.30, adj.p 1.20E-07), taurine (FC − 1.36, adj.p. 1.08E-06), N-acetyl-L-carnosine (FC −1.77, adj.p 8.20E-05), pyruvate (FC 1.60, adj.p 0.00337), succinate (FC −1.37, adj.p 0.0127). Interestingly, many of the identified metabolites suggested muscle origin of pathogenesis.

Creatinine, the secreted breakdown product of creatine, has been found to correlate with disease severity in SMA and spinal and bulbar muscular atrophy (SBMA) (22,23). Our metabolite profiling indicated that creatinine was decreased also in SMAJ (Fig 2B), whereas creatine was increased (Fig 2C). We validated the metabolite profiling result by measuring creatinine levels in an accredited laboratory and found that the two measurements correlated well (r = 0.74, p < 0.001). Similarly to CK, creatinine levels did not correlate with the symptom score (r = −0.11, p = 0.52) (Fig 2E). However, creatine levels correlated with the symptom score (r = 0.43, p = 0.009) (Fig 2F), and also with CK (r = 0.34, p = 0.03) (Fig 2G). We also detected declined levels of N-acetyl-L-carnosine (Fig 2H) and taurine in SMAJ patients’ serum (Fig 2I). These changes are likely to reflect alterations of muscle origin in SMAJ, and indicate muscle damage as well as decreased muscle mass.

### Indications of altered mitochondrial metabolism in SMAJ

Metabolite profiling showed elevated levels of pyruvate (Fig 3A), whereas succinate was decreased in SMAJ in comparison to controls (Fig 3B), indicating alterations in mitochondrial metabolism. Interestingly, however, lactate was not changed between the two groups (p = 0.34) (Fig 3C). These findings suggested that the mitochondrial origin of SMAJ contributed to the serum metabolite profile in patients. Thus, we tested if the previously characterized serum biomarkers of mitochondrial myopathy, FGF-21 or GDF-15 (24,25), were increased in SMAJ. However, we did not find the level of either marker to differ between patient and control groups (FGF-21 SMAJ median 162 pg/ml, controls 90 pg/m, [p = 0.15]; GDF-15 SMAJ median 653 pg/ml, controls 574 pg/ml, [p = 0.29]) (Fig 3D, E). None of the mitochondrial markers correlated with symptom score.

**Figure 3.**
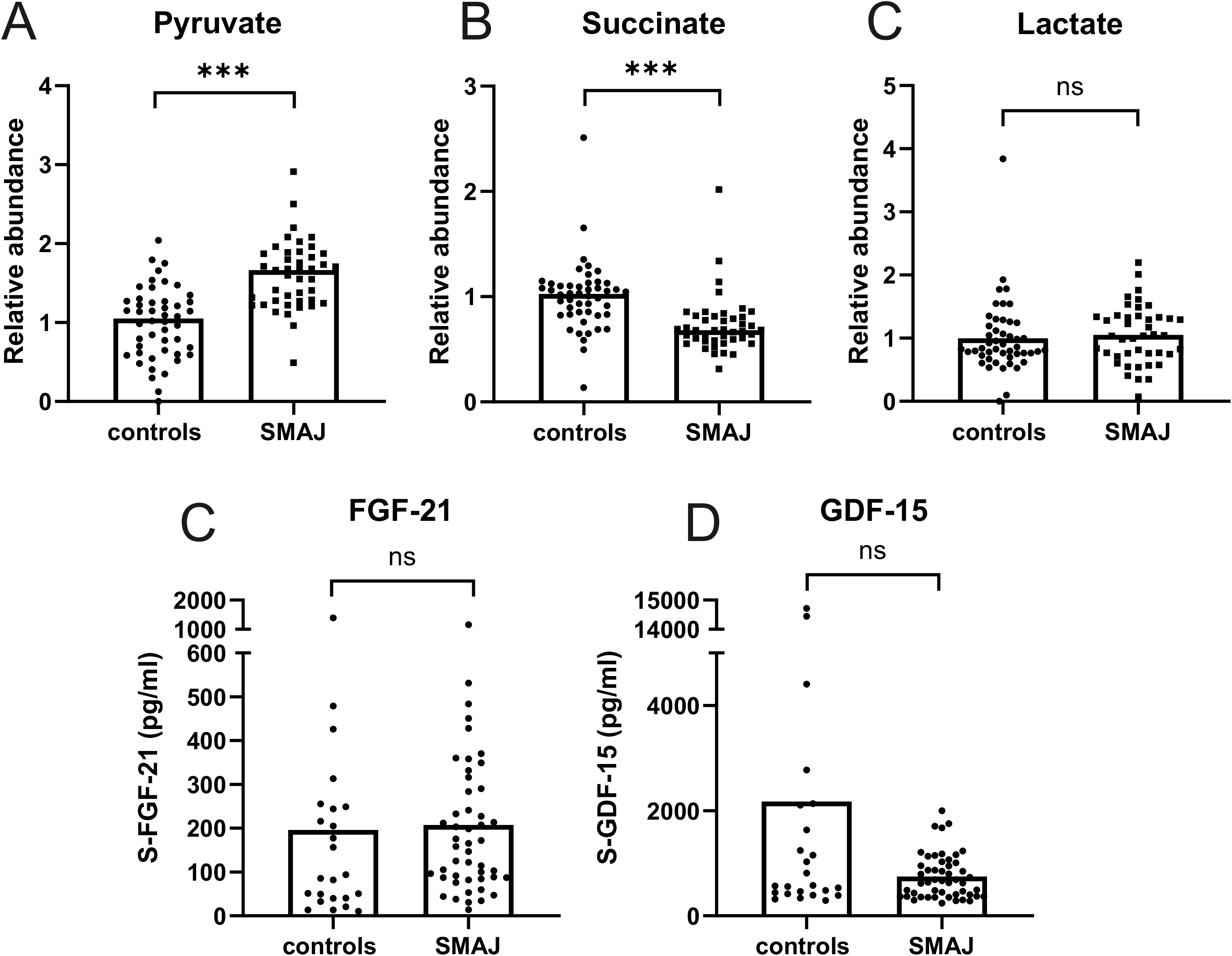
Mitochondrial disease biomarkers in SMAJ. Comparison of serum **A)** pyruvate, **B)** succinate, and **C)** lactate values between SMAJ patients (n=43) and controls (n=48) measured with mass spectrometer. Comparison of serum **D)** FGF-21 and **E)** GDF-15 levels between SMAJ patients (n=49) and controls (n=24) measured by ELISA assays. Median per group is indicated by bar. *** = p< 0.0001, Mann-Whitney U-test.

## DISCUSSION

Serum biomarkers are required as measures of treatment benefit in clinical trials of rare neurological diseases, which are challenged by small numbers of patients, and genetic and phenotypic heterogeneity. In addition, serum markers reveal features of pathogenesis, tissue involvement, and progression. Here we aimed at serum biomarker characterization in a unique homogeneous cohort of SMAJ patients with the identical pathogenic *CHCHD10* variant G66V, a founder mutation in Finland (26), surpassing ALS in prevalence in certain regions of the country (27). Our study resulted in the following main findings: 1) Unlike in several other motor neuron diseases, the neurodegeneration serum biomarkers NfL and GFAP are not elevated in SMAJ patients. 2) Serum metabolites of muscle-origin suggest that skeletal muscle pathogenesis is a major contributor to SMAJ. Of these, serum creatine correlated with symptom severity in SMAJ. 3) Abnormal CHCHD10 alters mitochondrial metabolite levels in serum (Fig 4).

**Figure 4.**
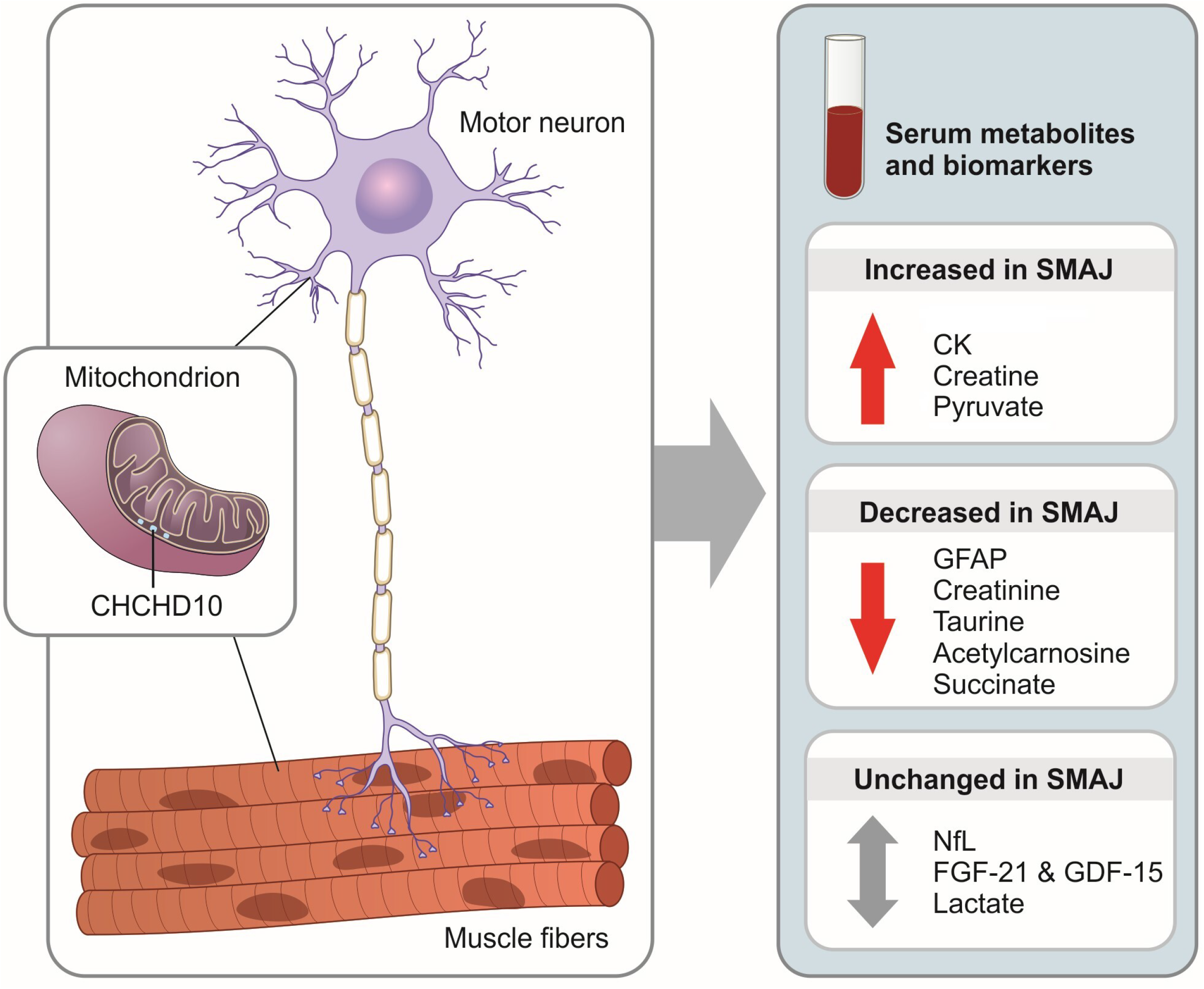
Summary of the SMAJ serum analyses of this study. SMAJ is caused by a dominant mutation in a gene, which codes for CHCHD10 protein located in mitochondrial intermembrane space. CHCHD10 defects impair mitochondrial respiration by an unknown mechanism. Serum analyses of biomarkers and metabolites suggest that SMAJ pathogenesis originates from skeletal muscle, showing altered muscle damage and mass markers, and signs of a metabolic shift in response to mitochondrial dysfunction.

Clear evidence of neurogenic muscle injury in SMAJ has been shown in EMG measurements as well as in muscle biopsies (1). Moreover, fasciculations indicate a neurogenic defect. Nevertheless, unlike in many neurodegenerative diseases, symptomatic SMAJ patients did not have an elevation of serum NfL despite of having a chronic injury process of spinal motor neurons (1). This contrasts with the high levels of neurofilaments, both NfL and pNFH (phosphorylated neurofilament heavy), in serum and CSF of patients with ALS (28,29), and in infants and young children with recently diagnosed 5q SMA, being also responsive to Nusinersen treatment (30,31). On the other hand, adults and adolescents with SMA disease did not have consistently increased NfL in CSF (32,33,34). As such, active ongoing motor neuron damage and broad anatomical involvement may be needed to produce a significant leakage of neurofilaments in a peripheral nervous system disease. The fact that SMAJ is slowly progressive and largely restricted to lower limbs may thus mean that the number of actively degenerating spinal motor neurons is not sufficient to elevate serum NfL. However, this hypothesis is in contrast with the elevation of serum NfL in patients with inherited peripheral neuropathies, including CMT1, CMT2 and hereditary sensory autonomic neuropathy (35).

An alternative explanation for the lack of NfL elevation among SMAJ patients is that the pathophysiology of SMAJ differs from neuropathies. An interesting parallel is provided by SBMA, in which the clinical picture includes slowly progressive weakness and wasting of bulbar and limb muscles (36). Similarly to SMAJ, in SBMA serum NfL was normal, but CK and creatine were increased whereas creatinine was decreased (22,37). These findings suggested that neurodegeneration in SBMA is a muscle-driven process (38). Thus it is possible that the primary cause of neuronal injury in SMAJ is not in the spinal motor neuron itself but driven non-cell autonomously by abnormalities in neuromuscular junction or muscle. Indeed, myopathic changes on muscle biopsy are more prominent in SMAJ and SBMA than in ALS (3). In addition to creatinine, decreased levels of taurine and N-acetyl-L-carnosine may also be indicators of decreased muscle mass in SMAJ patients. CK did not correlate to the clinical severity in SBMA or ALS (22), but creatinine predicted the severity in both SBMA and SMA, particularly the types 1 and 2 of the latter (22,23). Creatinine also correlated to therapeutic outcome of Nusinersen in SMA (39). However, only creatine correlated with the disease severity in SMAJ.

The absence of GFAP increase in SMAJ serum suggests that astrocytic injury or activation is not an important part of the disease process; the significantly lower levels of GFAP in SMAJ patients compared with controls may suggest impaired astrocytic activation but more studies are needed before conclusions regarding this can be drawn.

Altered pyruvate has been previously linked to primary mitochondrial myopathies (40,41). Increased pyruvate together with decreased succinate suggest a block in the TCA cycle, and thus glycolysis may be upregulated. The altered metabolites may thus be directly involved in compensatory pathways for balancing an energy deficiency in SMAJ. Interestingly, a recent study of ALS-linked *CHCHD10* variant reported similar findings for pyruvate and succinate in patient fibroblasts (14). The same study, as well as Chchd10 mouse models have indicated upregulation of ISR as part of pathogenesis, which leads to increased expression of muscle-secreted FGF-21 and GDF-15 (42). However, here we have shown that in the serum of patients with CHCHD10-linked disease, these growth factors are not increased. It is, however, possible that since p.G66V produces a milder phenotype than the other *CHCHD10* mutations, it does not initiate the ISR.

As a limitation of this study, although a large proportion of all SMAJ patients were included in the study, number of analyzed samples was still relatively small for a biomarker study. In addition, the symptom scoring system that we used was quite coarse, consisting of only three measurable sections of retrospectively collected data.

In conclusion, our study has established new insights into the pathophysiology of SMAJ in human patients and identified disease-associated metabolites, which may be beneficial in monitoring of treatment trials. Normal serum NfL may be clinically useful to differentiate SMAJ from early-stage ALS. Our findings on skeletal muscle may be relevant in the future studies of SMAJ and suggest that therapies designed for primary mitochondrial diseases may be worth testing also in SMAJ.

## Data Availability

Data availability statement Anonymized data will be shared by request from any qualified investigator.

## Acknowledgments

The authors thank the patients involved in the study and their families for participation and support. Riitta Lehtinen is acknowledged for assistance in ELISA measurements. We appreciate Biocentre Finland and Biocentre Kuopio for supporting LC-MS laboratory facility.

## Contributors

Study concept: JJ, SH, EP, MA, MJ, EY, HT. Data collection: JJ. Analysis and interpretation of data: JJ, SH, EY, HT. Statistical analysis: JK. NfL and GFAP measurements: HZ. Metabolite profiling: JK, ML. Patient data collection: AMS, MJ. Drafting of manuscript: JJ, SH, EY, HT. Critical revision of the manuscript: all authors.

